# Novel insight into the etiology of ischemic stroke gained by integrative transcriptome-wide association study

**DOI:** 10.1101/2023.03.30.23287918

**Authors:** Junghyun Jung, Zeyun Lu, Adam de Smith, Nicholas Mancuso

## Abstract

Stroke, characterized by sudden neurological deficits, is the second leading cause of death worldwide. Although genome-wide association studies (GWAS) have successfully identified many genomic regions associated with ischemic stroke (IS), the genes underlying risk and their regulatory mechanisms remain elusive. Here, we integrate a large-scale GWAS (N=1,296,908) for IS together with mRNA, splicing, enhancer RNA (eRNA) and protein expression data (N=11,588) from 50 tissues. We identify 136 genes/eRNA/proteins associated with IS risk across 54 independent genomic regions and find IS risk is most enriched for eQTLs in arterial and brain-related tissues. Focusing on IS-relevant tissues, we prioritize 9 genes/proteins using probabilistic fine-mapping TWAS analyses. In addition, we discover that blood cell traits, particularly reticulocyte cells, have shared genetic contributions with IS using TWAS-based pheWAS and genetic correlation analysis. Lastly, we integrate our findings with a large-scale pharmacological database and identify a secondary bile acid, deoxycholic acid, as a potential therapeutic component. Our work highlights IS risk genes/splicing-sites/enhancer activity/proteins with their phenotypic consequences using relevant tissues as well as identify potential therapeutic candidates for IS.

## Introduction

Stroke is a complex disease resulting from an interruption of blood flow to the brain [1]. A common type of stroke is ischemic stroke (IS), which is caused by cerebral infarction [2]. Diabetes, obesity, hypertension, and coronary artery disease are well-known risk factors for stroke [3–6], but the pathogenesis of IS is still largely unknown. Although genome-wide association studies (GWAS) have successfully identified genomic regions associated with IS outcomes, the genes underlying IS risk and their regulatory mechanisms remain elusive as the majority of associated variants are non-coding in nature [7,8].

Recently, the transcriptome-wide association study (TWAS) approach attempts to mitigate this gap in understanding by integrating GWAS associations together with molecular quantitative trait loci (molQTL) data [9,10]. Previous works have leveraged TWAS to identify candidate susceptibility genes for IS risk, however these analyses have three primary limitations. First, previous analyses were limited to integration of molQTLs measured in whole blood, adipose, and brain tissues [11,12], which may miss disease mechanisms in less understood or unknown disease-relevant tissues [13–15]. Second, prior works focused on integration of expression QTL (eQTL) and protein QTL (pQTL) [12,16], which may miss independent regulatory mechanisms important for IS risk. For example, an essential mechanism of gene regulation and a significant factor in genetic risk of disease is the genetic control of alternative splicing (i.e., sQTLs) [17]. Moreover, recent work demonstrated enhancers undergo activity-dependent transcription, resulting in the production of noncoding enhancer RNAs (eRNAs) which serve as a crucial hallmark of enhancer activation [18,19]. Third, while the GAGASTROKE prioritized relevant tissues for IS by leveraging eQTL data, it relied on the GTEx v7 study (N=388), [12], which had a smaller sample size than the European GTEx v8 study (N=588). Lastly, the L1000 Connectivity Map (CMap), a public database for large pharmalogical datasets, provides extensive gene expression profiles of thousands of compounds in various human cell lines [20]. Recent studies have successfully provided novel therapeutic candidates by utilizing TWAS results with the pharmalogical database [21,22].

Here, we integrate large-scale IS GWAS data (N=62,100 cases and 1,234,808 controls) with gene expression, alternative splicing, eRNA and protein abundance data (N=11,588) from 50 different tissues to identify IS susceptibility genes, tissues, and drug targets. We identify 136 genes/splicing sites/eRNA/proteins across 54 genomic regions whose genetically predicted activity is associated with IS risk using a multi-tissue mRNA/splicing/eRNA/protein transcriptome-wide association study (TWAS/spTWAS/eTWAS/PWAS). We leverage TWAS results to identify tissues relevant for IS risk and find arterial and brain most enriched for eQTL mediated heritability. Focusing on the IS-relevant tissues, we perform probabilistic fine-mapping analyses of TWAS results to prioritize 9 putative causal genes/proteins. Among them, only 3 genes/proteins (i.e., *FOX2, F11, MMP12*) were identified in previous GWAS or TWAS studies [7,12,16]. In addition, we conduct a TWAS-based pheWAS analysis to understand the phenotypic consequences of identified IS susceptibility genes, and the identified reticulocyte cell traits are significantly correlated with IS. In addition, we discover that reticulocyte traits have shared genetic contributions with IS using TWAS-based pheWAS and genetic correlation analysis. Lastly, to find therapeutic drug candidates for IS risk, we integrate our TWAS findings with 308,872 pairs of compound and compound-perturbed cell-type specific gene expression alterations from the L1000 Connectivity Map. Using this approach, we detect a secondary bile acid, deoxycholic-acid (DCA), as a potential therapeutic component. Overall, our results shed light on underlying molecular mechanisms and tissue contexts that cause ischemic stroke.

## Materials and Methods

### Ischemic stroke GWAS summary statistics

IS GWAS summary statistics from GIGASTROKE consortium [16] were downloaded from the GWAS Catalog (GCST90104540) (see **URLs**). We restricted summary statistics data to ischemic stroke (AIS) results from 1,296,908 individuals of predominantly European ancestry (62,100 cases and 1,234,808 controls). Next, we filtered summary statistics data to exclude SNPs with minor allele frequency (MAF) < 0.01 or any SNPs with strand-ambiguous variants (i.e. A/T or C/G; or vice-versa) using the focus munge tool [23], resulting in 6,335,571 bi-allelic SNPs for downstream analyses.

### Reference functional data for predictive models of eQTL/spQTL/EeQLT/pQTL

To perform TWAS and spTWAS, we generated predictive models of gene and splicing expression using individuals from the Genotype-Tissue Expression Project (GTEx) v8 [24] using a modified FUSION script (see **URLs**). We downloaded genotype, phenotype, and covariate information from European-American subjects in the GTEx v8 study (48 tissues; N□=588). We defined the *cis*-mapping window as ±500kb around the transcription start site (TSS) after filtering based on minor allele frequency (MAF) < 0.005, Hardy-Weinberg Equilibrium (HWE) < 1 × 10^−5^. We included the reference set of covariates from the GTEx v8 eQTL/spQTL analyses, which included first five genotype principal components (PCs), 15 hidden covariate derived from Probabilistic Estimation of Expression Residuals (PEER) [25] factors, whole genome sequencing (WGS) platform (HiSeq 2000 or HiSeq X), WGS library preparation protocol (PCR-based or PCR-free), and donor gender. We estimated *cis*-SNP heritability 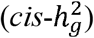 of each model using REML as implemented in Genome-wide Complex Trait Analysis (GCTA) [26]. We focused on genes and splicing sites with nominally significant estimates of 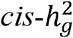 (*p*-value < 0.01), which resulted in 293,295 total tissue-gene pairs from 27,549 unique genes and 371,441 total splicing sites from 16,871 unique genes. To train and generate predictive models, we fitted LASSO [27], Elastic Net [28], and the Sum of Single Effects model (SuSiE) [29]. The best performing model for each gene/tissue (splice-site/tissue) context was selected by calculating cross-validation prediction accuracy, as implemented in the FUSION pipeline.

To perform enhancer TWAS analysis (eTWAS), we downloaded precalculated FUSION weights for enhancer eQTL (EeQTL) data from two brain tissues, the dorsolateral prefrontal cortex (DLPFC) (N=486) and the anterior cingulate cortex (ACC) (N=402), in CommonMind Consortium (CMC) [19]. Briefly, the LASSO [27], Elastic Net [28] were used to train and generate the eTWAS predictive models for the ACC (4,907 predictive models) and DLPFC (5,715 predictive models). In addition, we also downloaded fitted prediction models of gene expression in both ACC (8,021 predictive models) and DLPFC (8,946 predictive models) tissues. For detailed information of the eTWAS and TWAS predicted models, see the previously described ref [19].

To perform a proteome-wide association study (PWAS), we downloaded fitted prediction models of protein abundance trained using individuals from the INTERVAL (N=3,301) [30] and Atherosclerosis Risk in Communities (ARIC) [31] cohorts (see **URLs**). In the ARIC cohort, we focused only on the European ancestry dataset (N=7,213). Briefly, to train and generate the PWAS predictive models using FUSION script, the LASSO [27], Elastic Net [28], and SuSiE [29] were used for the INTERVAL (994 predictive models) and the Elastic Net [28] was used for the ARIC (1,305 predictive models). For detailed information of the PWAS predicted models, see the previously described INTERVAL [32] and the ARIC [33].

### Transcriptome-wide association study analyses

We performed TWAS, spTWAS, eTWAS, or PWAS analyses using FUSION [9] with the trained GTExv8 [24], CMC [19], or INTERVAL [32] and ARIC [33] models, respectively. European LD reference data from the 1000G project [34] was used for TWAS analysis. We excluded TWAS associations from human leukocyte antigen (HLA) regions due to the complex LD patterns. The significance threshold of TWAS associations was determined using a per-tissue Bonferroni correction (Avg num tests = 6770; see **Supplementary Table 1**). We then carried out an adaptive permutation test using TWAS test statistics for each tissue panel. For this analysis, 10^6^ maximum number of permutations was used, and the significance threshold was corrected with per-tissue Bonferroni correction.

To provide partial support for findings from our FUSION TWAS analyses, we leveraged S-PrediXcan [35] with pre-trained Multivariate Adaptive Shrinkage in R (MASHR)-based models of PredictDB [36,37]. In GTEx v8, we tested 648,028 and 1,681,295 models of expression and splicing site, respectively.

### Colocalization analysis

We performed colocalization analysis to test whether the same causal variants were shared between stroke GWAS and gene/protein expression levels. We used coloc r package [29] together with stroke GWAS summary statistics and marginal molecular QTL (molQTL) results from FUSION prediction models. The evidence of colocalization was defined as PP_3_ + PP_4_ ≥ 0.8 and PP_4_/PP_3_ ≥ 2 where posterior probability (PP) was obtained from hypothesis *H*_3_ and *H*_4_. *H*_3_ is the PP of GWAS and eQTL signals are associated with different causal variants, and *H*_*4*_ is the PP of GWAS and eQTL signals are associated and share a single causal variant.

### Mediated Expression Score regression analyses

To identify tissues relevant for IS risk, we used Mediated Expression Score Regression (MESC) to estimate the proportion of heritability mediated by assayed gene expression levels 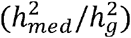 [38]. For MESC expression scores, the eQTL effect sizes using the SuSiE model [29] and expression *cis*-heritability using REML were imported from FUSION GTEx v8 weights. Only SNPs from the HapMap3 [39] were kept for this research.

### Fine-mapping of TWAS associations

To differentiate between causal and tagging associations at TWAS risk regions, we performed probabilistic fine-mapping of TWAS results to prioritize genes using the tool FOCUS [23]. We generated an eQTL/spQTL/pQTL weight database for FOCUS by importing our trained GTEx v8 weights from FUSION. To determine approximately independent genomic regions, we used LD block architecture GRCh19 provided by Berisa and Pickrell *et al*. [40] (see **URLs**).

### Phenome-wide association studies and genetic correlation analyses

To understand the phenotypic consequences of identified TWAS/spTWAS/PWAS associations, we performed a phenome-wide association study (pheWAS) for each identified gene using PhenomeXcan [41]. Using the TWAS/spTWAS/PWAS genes, phenotypes from the results of pheWAS analysis were reported based on transcriptome-wide significance (P-value < 2.25 × 10-^6^). In addition, the PheWAS result was filtered by traits involving at least five TWAS genes.

To determine the genetic relationship between IS and the phenotypes identified from PhenomeXcan, we performed genome-wide genetic correlation analyses using RHOGE [42] with publicly available GWAS summary statistics from the UK Biobank (see **URLs**). First, TWAS/spTWAS/PWAS analyses were performed using the same FUSION pipeline in the IS analysis. Next, we estimated the genome-wide genetic correlation between the IS and the PhenomeXcan traits derived from TWAS analysis using LD block architecture GRCh38 (see **URLs**) MacDonald *et al*. [43]. In addition, we performed SNP-based genetic correlation analysis using LD Score Regression (LDSC) [44] with the GWAS summary statistics data.

### Mendelian randomization analysis

To assess causal effects of reticulocyte traits on IS risk, we performed Mendelian Randomization (MR) analysis. To extract instruments for use in MR from the 5 reticulocyte traits including “Reticulocyte percentage (ukb-d-30240_irnt)”, “Reticulocyte count (ukb-d-30250_irnt)”, “Immature reticulocyte fraction (ukb-d-30280_irnt)”, “High light scatter reticulocyte percentage (ukb-d-30290_irnt)”, and “High light scatter reticulocyte count (ukb-d-30300_irnt)”, we leveraged extract_instruments function in TwoSampleMR r package [45]. In addition, For the reverse relationship between reticulocyte traits and IS risk, the fine-mapping results of GIGASTROKE [16] were used as instruments. After harmonizing the effect of a SNP on the between IS and reticulocyte traits to the same allele, the MR analysis was conducted using 5 MR method (“MR Egger”, “Weighted median”, “Inverse variance weighted”, “Simple mode”, and “Weighted mode”).

### Drug repositioning analysis

To identify potential drug candidates for IS, we used Trans-Phar [21] by comparing the inverse expression profiles between genetically regulated gene expression from TWAS with compound-induced gene expression profiles from a large-scale pharmacological database. Briefly, we obtained differentially expressed genes (DEGs) data from ref. [21] for each compound-induced gene expression data and used 13 tissue/cell-type categories assigned for 29 GTEx tissues and 77 L1000 Connectivity Map (CMap) cell types [21]. Then, we computed the rank correlations using Spearman’s rank correlation coefficient between the top 10% TWAS genes from each GTEx tissue and each CMAP expression level in the same group of tissues or cell types. In addition, we also calculated the Spearman’s rank correlation coefficient using the 134 TWAS/spTWAS/PWAS genes. Finally, a total of 308,872 P-values were collected for each correlation analysis. The significance threshold of the inverse correlation analysis was determined using a per-tissue or/and per-cell type Bonferroni correction (see **Supplementary Table 2**).

## Results

### Multi-tissue TWAS/spTWAS/eTWAS/PWAS identifies genes/proteins associated with IS risk

To identify susceptibility genes and proteins for IS risk, we conducted a multi-tissue TWAS, spTWAS, and PWAS analysis by integrating large-scale IS GWAS summary statistics (N=1,296,908) together with eQTL/spQTL data from GTEx v8 [24] in addition to plasma pQTL data from the INTERVAL [30] and the ARIC [33] studies. Moreover, we performed enhancer TWAS analysis (eTWAS) using brain EeQTL from CMC [19] to investigate the impact of genetic regulation of expressed enhancers on IS risk (see **Methods**). Focusing on predicted mRNA expression, we tested 309,826 panel-specific expression models across 28,073 genes and identified 268 TWAS associations. Significant associations represented 84 genes across 50 tissues and 41 independent 1 Mb genomic regions based on a per-panel Bonferroni correction threshold (see **Fig. 1A** and **Supplementary Table 3**). Next, to shed light on the role of alternative splicing for IS risk, we performed a multi-tissue splicing transcriptome-wide association study (spTWAS; see Methods). Of the 370,815 panel-specific splicing models with 16,848 tested genes, we identified 498 spTWAS associations. Associations represented 69 unique genes across 48 tissues at 29 independent 1Mb genomic regions (see **Fig. 1B** and **Supplementary Table 4**). In addition, we carried out an eTWAS to investigate expressed enhancer effects in two brain tissues using 10,622 tissue-specific eTWAS models with 8,397 eRNAs. We identified two enhancers, chr4:186651582:186652095 and chr16:87541308:87541807, in DLPFC and ACC tissues, respectively (see **Fig. 1C** and **Supplementary Table 5**). Lastly, we focused on the impact of genetically predicted protein abundance by performing a PWAS using 2,299 panel-specific protein abundance models with 1,556 proteins. We identified 7 PWAS associations across 6 proteins with 6 independent 1 Mb regions (see **Fig. 1D** and **Supplementary Table 6**). PWAS results from both pQTL panels showed strong positive correlation (R = 0.79; P-value < 2.2 × 10^−16^; see **Fig. S1**), suggesting that the genomic component of protein abundance is well-captured by fitted models. Comparing the analysis results of TWAS and spTWAS, a total of 21 genes were implicated by both approaches, with *ABO and F11* representing the genes or proteins identified by all three approaches (see **Fig. S2**). In addition, we identified 54 genomic regions with a total of 136 susceptibility genes, splicing sites, eRNA, and proteins based on multi-tissue TWAS/spTWAS/PWAS approaches. Among the genomic regions, 20 genomic regions have been implicated in GWAS analysis [16].

**Figure 1.**
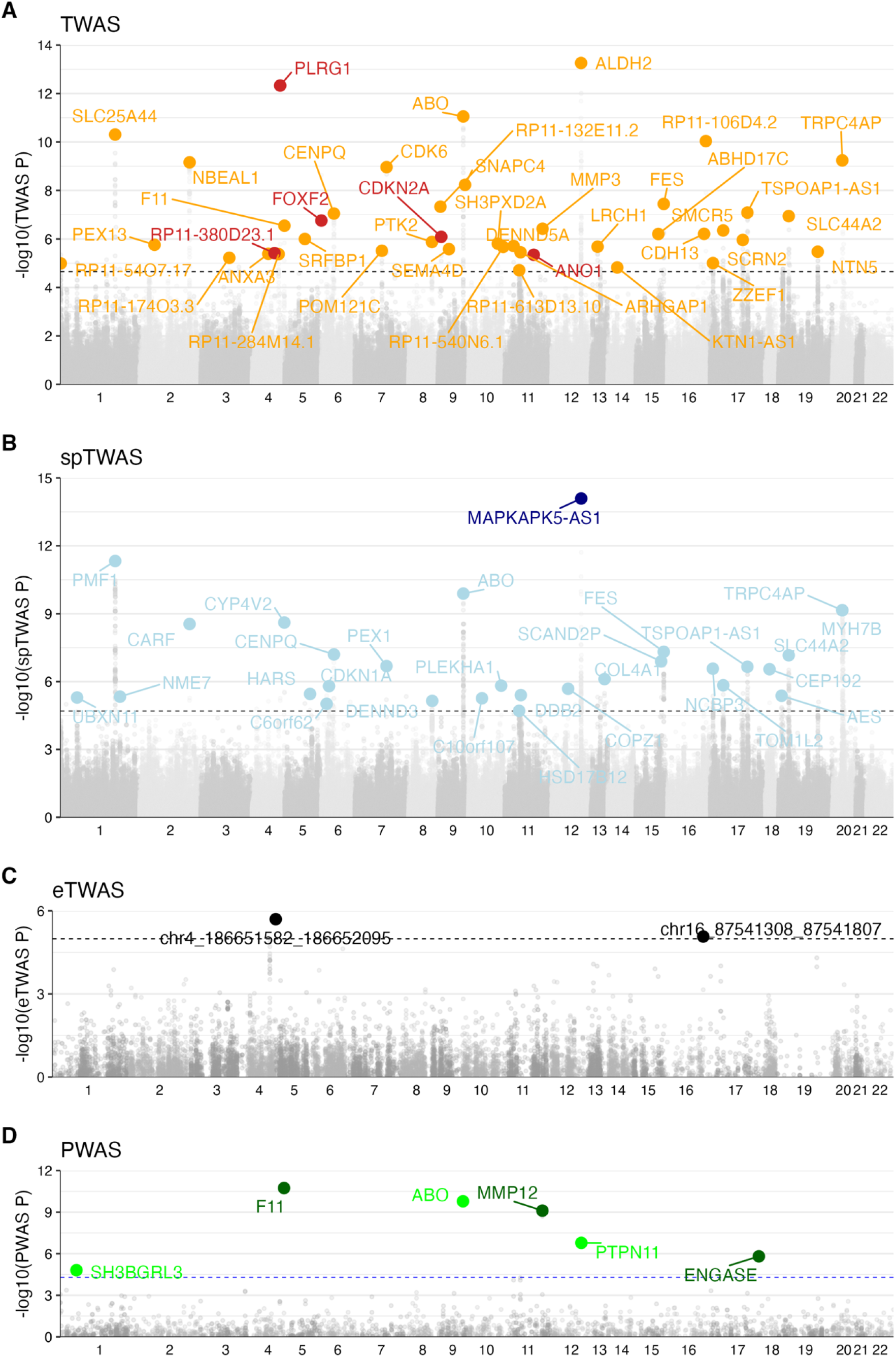
Multi-tissue TWAS/spTWAS/PWAS for IS risk. Manhattan plots of multi-tissue (A) TWAS, (B) spTWAS, (C) eTWAS, and (D) PWAS. Each point corresponds to a P-value (y-axis) of TWAS/spTWAS/eTWAS/PWAS associations across reference panels and chromosomes (x-axis). The most significant associations of TWAS/spTWAS/eTWAS/PWAS genes among reference panels represent orange, light blue, black and green, respectively. The black dotted lines represent the maximum significant thresholds in this multi-tissue analysis (**Supplementary Table 1**). The thresholds of TWAS/spTWAS/eTWAS/PWAS indicate 2.23 × 10^−5^, 2.00 × 10^−5^, 1.02 × 10^−5^, and 5.03 × 10^−5^, respectively. The putative causal genes (PIP > 0.8) in **Table 1** represented gene/proteins of TWAS/spTWAS/PWAS indicate red, navy, and dark green, respectively.

To provide additional support for genes identified using TWAS, we re-performed analyses using independent approaches and prediction models. First, we compared TWAS and spTWAS results with those computed by S-PrediXcan [35], an independent method to perform a TWAS analysis and fit predictive models. We found S-PrediXcan results were strongly correlated with FUSION-based TWAS (R = 0.80; P-value < 2.2 × 10^−16^) and spTWAS (R = 0.82; P-value < 2.2 × 10^−16^; see **Fig. S3**). Second, we carried out a co-localization analysis, which reports the posterior evidence that two phenotypes share a causal variant. Here, we identified 51.49% (138 out of 268) TWAS associations exhibited evidence of co-localization between GWAS signals and eQTLs (see **Supplementary Table 3**). Similarly, among the 498 spTWAS associations, we observed that 271 associations (54.41%) had evidence of co-localization between IS GWAS signal and spQTL association (see **Supplementary Table 4**). Moreover, we observed that the *H*_4_ posterior probability of co-localization, correlated with the TWAS (R = 0.52; P-value < 2.2 × 10^−16^) and spTWAS (R = 0.54; P-value < 2.2 × 10^−16^) (see **Fig. S4**). Repeating this analysis for PWAS signals, we observed *ABO*, F11, and *MMP12* displayed evidence of colocalization between GWAS and pQTL signals (see **Supplementary Table 6**). Despite eTWAS identifying 2 eTWAS signals, we found little support for colocalization between eTWAS and IS GWAS (see **Supplementary Table 5**). The GIGASTROKE consortium recently performed IS TWAS analyses based on GTEx v7 prediction models and identified 17 genes across brain, artery, and heart tissues [16]. We sought to assess the stability of these associations by comparing them with our results which leveraged the larger GTEx v8 and INTERVAL/ARIC cohorts. Among the 17 TWAS genes identified in the previous analyses, we found 7 replicated in our TWAS results (average Bonferroni across tissues, P < 7.39 × 10^−6^). Similarly, we observed a significant correlation between the TWAS effect sizes in the original GIGASTROKE study with those computed using our prediction models (R = 0.73; see **Fig. S5**). In summary, our findings support a role for genetically regulated expression, splicing, and proteome levels contributing IS risk.

### Relevant tissues for IS risk include brain and arterial tissues

Given the broad number of tissues exhibiting TWAS/spTWAS/PWAS associations, we sought to quantify which tissues are most relevant for IS risk for each molecular context. Specifically, we estimated the proportion of heritability mediated by *cis*-QTL of gene expression levels 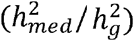 and alternative splicing levels 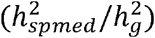 using mediated expression score regression (MESC; see **Methods**) [38]. Using this approach, we identified 3 tissues which exhibited 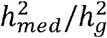 estimates greater than 0 at nominal significance (P-value < 0.05). We found “Artery - Aorta” exhibited the greatest 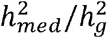 value (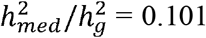, s.e= 0.038; P-value = 4.55 × 10^−3^), followed by “Esophagus_Muscularis” (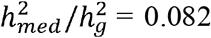, s.e= 0.040; P-value = 1.86 × 10^−2^) and “Brain_Amygdala” (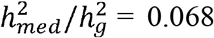, s.e= 0.040; P-value = 4.55 × 10^−2^) (see **Fig. 2**). Results across expression and splicing were consistent MESC across tissues (R = 0.33; P-value = 2.24 × 10^−2^; see **Fig. S6**). Previous studies reported that the risk of gastroesophageal reflux disease (GERD) is higher among stroke patients [46] and increases young adult stroke risk [47], suggesting that risk factors shared by individuals with GERD and stroke, such as diet, smoking, obesity, and metabolic syndrome, may account for this association. Our results are supportive of understood IS etiology in which IS occurs due to blood clotting or fatty deposits caused by atherosclerosis that obstruct an artery supplying blood to the brain [48]. Atherosclerosis is commonly thought of as a condition that affects the heart; however, it can also impact arteries located anywhere in the body [49]. In the splicing MESC, “Brain - Nucleus accumbens (basal ganglia)” (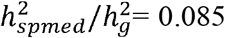 and s.e. = 0.056) was a high rank in the splicing MESC (see **Fig. S7**).

**Figure 2.**
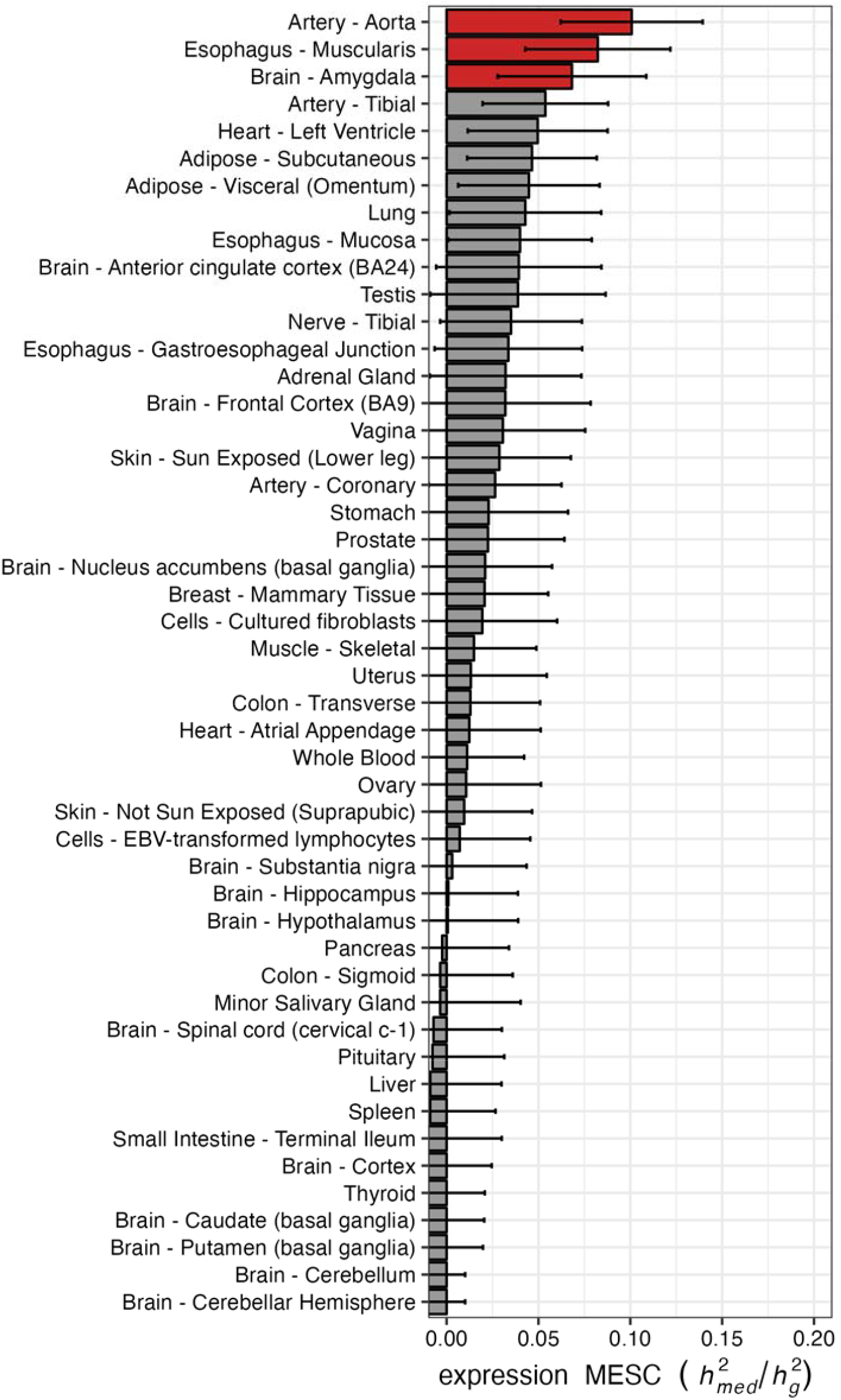
Proportion of heritability mediated by gene expression levels. The bar plots correspond the estimated expression MESC 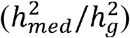 in each tissue panel in GTEx v8. Each error bar indicates jackknife standard errors. The red color represents significant tissue panels (P-value < 0.05).

### Fine-mapping analysis identifies 9 causal genes/proteins for IS risk

Next, we performed TWAS fine-mapping analysis to identify putative causal genes with multiple signals in the TWAS region prioritizing IS-associated tissues obtained from the MESC analysis [23] (Fig. 2). Among the 84 TWAS significant genes, we identified 5 with posterior inclusion probability (PIP) > 0.8, which we denote as putative causal genes for IS (see **Table 1**). For example, *ANO1* (PIP=0.89; also known as *TMEM16A*) is suggested to regulate calcium-activated chloride channels and has prior evidence in mouse ischemic stroke models where inhibition of *ANO1* expression levels attenuated ischemic brain injury by neurological impairment [50]. We separately performed spTWAS fine-mapping analysis to identify splice variation that may be causally related to IS risk prioritizing brain tissues obtained from splicing MESC analyses. Of the 69 spTWAS significant genes, we identified only *MAPKAPK5-AS1* with PIP > 0.8; see **Table 1**. The mitogen-activated protein kinase (MAPK)-activated protein kinase 5 (APK5), a member of the serine/threonine kinase family, is activated by cellular stress and proinflammatory cytokines [51]. The MAPKAPK5 Antisense RNA 1 (MAPKAPK5-AS1) prevents MAPKAPK5 from being translated into a protein and recent study showed that IS-like pathology was ameliorated by inhibiting the MAPK signaling pathway [52]. Lastly, we performed PWAS fine-mapping to identify protein levels causally relevant to IS risk. Of the 7 PWAS significant associations identified, we found 3 with PIP > 0.8 (see **Table 1**). Of these 3, F11 and MMP12 genes were identified in a previous TWAS of IS risk [12]. Together, we prioritize 9 putative causal genes/proteins based on relevant tissue for IS risk.

**Table 1.**
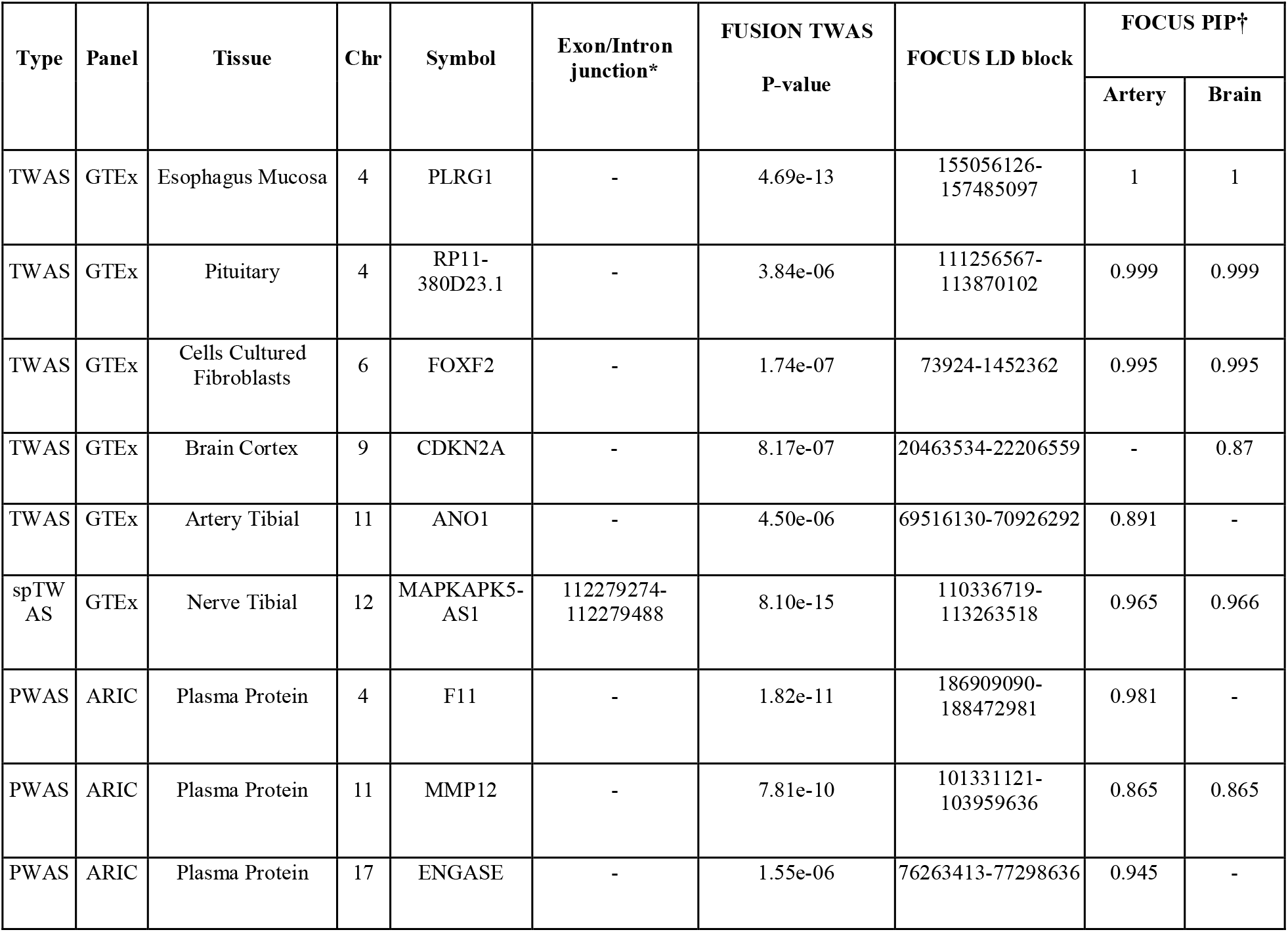
Putative causal genes/splicing site/proteins from TWAS/spTWAS/PWAS (PIP > 0.8). *: All genomic locations are GRCh37. †: FOCUS PIP values are obtained by tissue prioritization using “Artery” or “Brain” tissue.

### IS susceptibility genes correlate with reticulocyte cell traits

To understand the phenotypic consequences of identified IS susceptibility genes, we performed a TWAS-based pheWAS analysis (see **Methods**). Analogous to the TWAS approach, the TWAS-based pheWAS approach has produced more biologically interpretable results by mapping the genome to the phenome using the transcriptome [41]. We identified 71 traits across a broad range of physical measures (47.9%) and blood cell traits (33.8%) (see **Fig. S8**). In the category of physical measures, an average of 25.4% of genes (11.7/46 genes) detected only by spTWAS analysis were enriched, whereas an average of 34.7% of genes (7.3/21 genes) associated with both TWAS and spTWAS were mostly enriched in blood cell traits (see **Supplementary Table 7**). Next, we applied genetic correlation analysis to test whether the PheWAS traits shared genetic contributions with IS at a transcriptome- or proteome-wide level (see **Methods**). The results show that the well-known risk factors for stroke, such as “Diastolic blood pressure. automated reading(4079_irnt)”, “Systolic blood pressure, automated reading (4080_irnt)”, “Non-cancer illness code, self-reported: hypertension (20002_1065)”, “Vascular/heart problems diagnosed by doctor: High blood pressure (6150_4)”, and “Non-cancer illness code, self-reported: high cholesterol (20002_1473)” [4,53,54], were significantly correlated with IS in the physical measures and medical conditions categories (FDR < 0.05) (see **Fig. 3** and **Supplementary Table 8**). Notably, 5 reticulocyte traits, including “Reticulocyte count (30250_irnt)”, “Reticulocyte percentage (30240_irnt)”, “High light scatter reticulocyte percentage(30290_irnt)”, “High light scatter reticulocyte count(30300_irnt), and “Immature reticulocyte fraction (30280_irnt)”, were significantly positively correlated with IS in TWAS and spTWAS levels in the blood cell traits (FDR < 0.05). Reticulocytes are slightly immature red blood cells, and immature reticulocyte fraction levels are associated with acute infection, chronic renal insufficiency, and hematologic diseases [55]. However, we found little evidence for a causal relationship between IS risk and reticulocyte traits (see **Fig. S9**). Moreover, previous studies showed that high reticulocyte count or reticulocytosis are risk factors for stroke in children with sickle cell disease [56–58]. Additionally, we also used LD-score regression (LDSC) to measure shared genetic contributions between traits at the genome-wide level. We found significant correlations between IS and the reticulocyte traits and the well-known risk factors. Collectively, our results show that vascular traits and blood cell traits, especially reticulocyte cells, have shared genetic contributions with IS.

**Figure 3.**
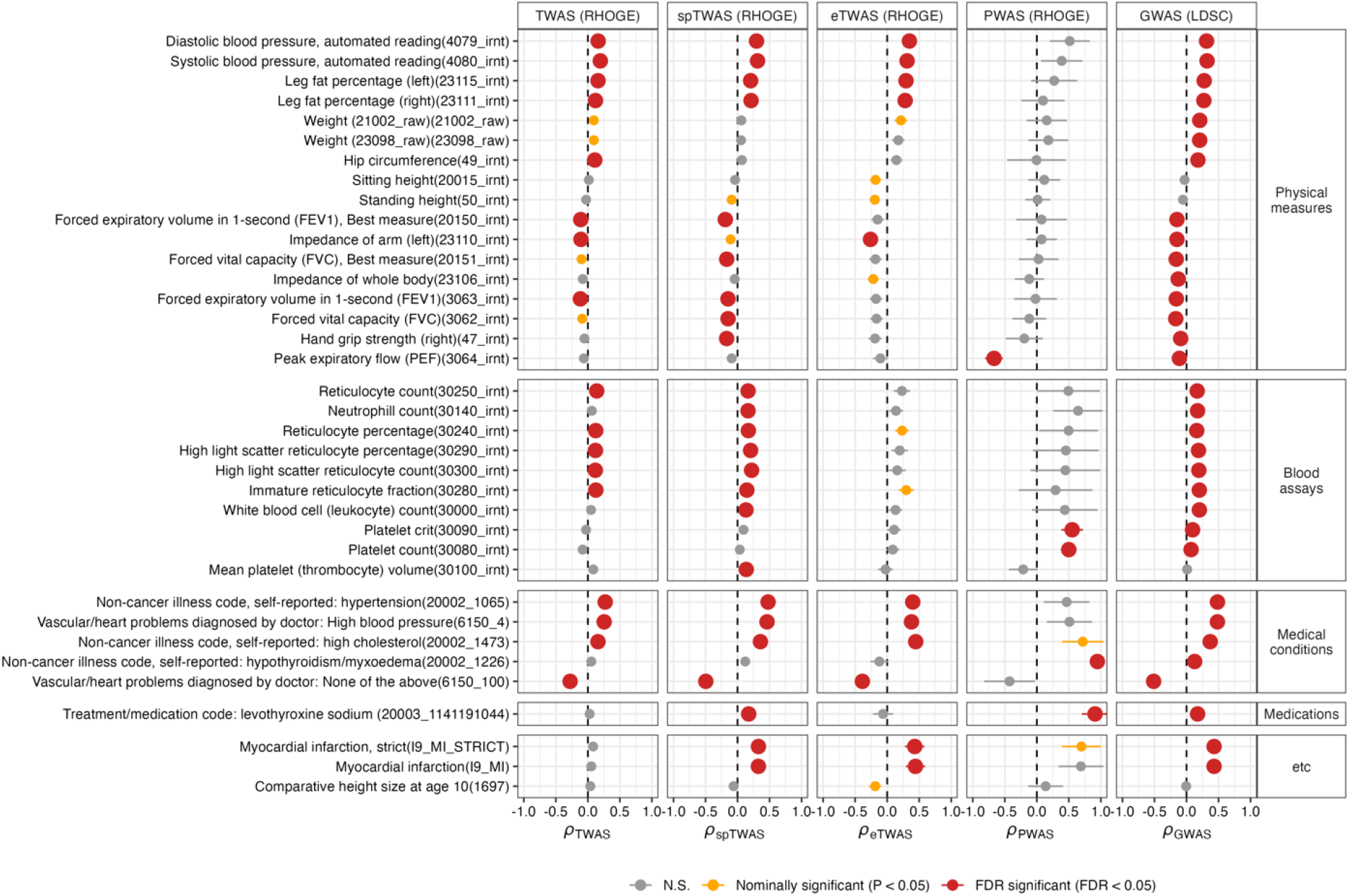
Genetic correlation between IS and UKBB phenotypes. Each point indicates a genetic correlation with standard errors from TWAS/spTWAS/eTWAS/PWAS. Significance levels are represented by red or orange color. We use the following abbreviations *ρ*_*TWAS*_, transcriptomic correlation analysis using TWAS; *ρ*_*spTWAS*_, transcriptomic correlation analysis using spTWAS; *ρ*_*eTWAS*_, transcriptomic correlation analysis using eTWAS; *ρ*_*PWAS*_, proteomic correlation analysis using PWAS; *ρ*_*GWAS*_, genomic correlation analysis using GWAS. In the figure, we show results indicating significance in at least one TWAS/spTWAS/eTWAS/PWAS analysis.

### TransPhar analysis identifies secondary bile acids as potential drug candidates for IS

Next, we sought to find potential drug candidates for IS treatment by evaluating an inverse expression relationship with compound-induced gene expression profiles from a large-scale pharmacological database, L1000 CMap [20]. We computed the rank correlations using Spearman’s rank correlation coefficient between the top 10% TWAS genes (or the 134 TWAS/spTWAS/PWAS genes) from each GTEx tissue (a total of 29 GTEx tissues) and each CMAP expression level in the same group of tissues or cell types. The top 10% TWAS genes of each tissue contained an average of about 22.26% of the TWAS/spTWAS/PWAS genes (mean 29.82 genes) among the 134 TWAS/spTWAS/PWAS genes or proteins (see **Fig. S10**). We obtained 308,872 relationships from all the tissue/cell type-compound pairs corresponding to each correlation analysis, and the results from the top 10% TWAS genes and the 134 TWAS/spTWAS/PWAS genes were significantly correlated in each tissue (see **Fig. S11**).

Finally, we found two significant TWAS-compound linkages based on false discovery rate (FDR) correction (see **Table 2, Fig. S12, and S13**). The two compounds were associated with brain tissue; among them, the mechanism of action has been elucidated for deoxycholic-acid (DCA). The DCA is classified as secondary bile acids, which are produced by gut microbiota [59]. Previous studies showed that other types of secondary bile acids, such as Ursodeoxycholic acid (UDCA) and tauroursodeoxycholic acid (TUDCA), have demonstrated neuroprotective effects in diverse models of neurodegenerative disorders [60–64] as well as stroke [65]. Notably, lower levels of total bile acid excretion is linked to an increased risk of stroke and death [66]. The GIGASTROKE consortium carried out the inverse relationship analysis between TWAS gene and compound expression levels using GTEx v7 data, but there were no significant results for IS [16]. In summary, our results suggest that a negative correlation between the TWAS genes and compound-induced gene expression levels provides the potential drug candidates for IS.

**Table 2.** Potential drug candidates for IS. Rho (ρ) represents Spearman’s Rank correlation coefficient, and FDR represents the false discovery rate.

| Tissue/cell-type categories | GTEx panel (tissue) | CMap L1000 cell line | CMap L1000 library (dose, h) | Mechanism of action | rho ($\rho$) | P-value | FDR |
| --- | --- | --- | --- | --- | --- | --- | --- |
| Brain | Brain - Frontal Cortex BA9 | Neural progenitor cells (NPC) | Deoxycholic-acid (DCA) (10μM, 24h) | Fat emulsification | -0.301 | 8.00e-06 | 5.46e-02 |
| | | | BRD-K41878610 (10μM, 24h) | NA | -0.293 | 1.32e-05 | 5.46e-02 |

## Discussion

Stroke is the second leading cause of death worldwide, and the main cause of stroke is ischemic stroke (IS) due to cerebral infarction [2]. In this work, we identify 136 susceptibility genes, splicing sites, eRNA, and proteins spanning 54 genomic regions based on multi-tissue TWAS/spTWAS/PWAS analysis. Among them, 34 genomic regions were not discovered by the original GWAS study. Consequently, we highlight 9 potential causal genes/proteins using probabilistic fine-mapping analysis of the TWAS results with a focus on the IS-relevant tissues, including aorta artery and brain estimated by a total fraction of disease heritability mediated by gene expression levels. Moreover, we discovered that blood cell traits, particularly reticulocyte cells, have shared genetic contributions with IS. Lastly, we detected 2 potential therapeutic compounds for using inverse expression profiles between TWAS results with compound-induced gene expression profiles from a large-scale pharmacological database.

The genes underlying IS risk and their regulatory mechanisms are still unknown, despite the fact that GWAS have effectively identified numerous genomic regions related with IS, because around 90% of the GWAS loci are located in non-coding regions [7,8]. However, the TWAS methodology has produced more biologically interpretable results by integrating GWAS results with mQTL data, such as eQTL, spQTL, EnQTL, and pQTL [9,10,19]. Consistent with the TWAS approach, the PhenomeXcan has allowed us to identify the mediating role of gene expression in complex traits and convert variant-phenotype associations into gene-phenotype associations by providing biological hypotheses [9,36,67].

A notable finding was the significantly positive correlation between IS risk and reticulocyte traits (see **Fig. 3**). Reticulocytes are slightly immature red blood cells, and higher reticulocyte counts or percentages indicate higher hemolysis, which ultimately results in an increase in the amount of cell-free hemoglobin (CFH) in the blood [68]. The previous studies showed that the hemolysis and the CFH increases inflammation [69] and mediate vascular damage [70]. The findings suggested that a shared causal mechanism influence both the high level of reticulocytes with hemolysis and risk of IS.

After an initial ischemic stroke or transient ischemic attack (TIA) which is a short episode in which temporary blockage of blood flow to the brain, the annual risk of future ischemic stroke is 3 to 5% [71]. The TIA does not leave an impairment, but affected individuals are at increased risk of future ischemic events, especially in the days and weeks immediately after symptoms resolve [72]. The prompt initiation of a coordinated preventive strategy for IS is essential [73].

Recent studies have reported that the probability of success for clinical trials whose therapeutic targets are supported by human genetic information is approximately twice the probability of success for unsupported projects [74,75]. Taking advantage of the rapid growth of genomics fields, the utilization of human genetics information for new therapeutics have been performed in recent years [21,22,76,77]. Importantly, 66% (33 out of 50) of the FDA-approved new drugs were supported by human genetic information in 2021 [78]. In addition to genetic data, recent gene expression datasets have been leveraged for drug repositioning [79,80]. In particular, recent studies have investigated the utility of TWAS in drug discovery or repositioning [21,22]. One primary advantage of the TWAS-based drug repositioning strategy, which integrates large-scale GWAS together with gene expression data, compared with datasets solely comprised of expression is a vastly larger sample size. In general, the sample sizes of GWAS are many orders of magnitude larger (e.g., tens to hundreds of thousands) than those of gene expression datasets (e.g., tens to hundreds). In addition, the TWAS approach enables identifying potential therapeutic compounds that affect multiple tissues (e.g., heart, brain) or cell type-specific contexts by leveraging independent reference eQTL data. On the other hand, the expression data related to the tissue of interest are not easily accessible for many diseases, including IS.

To shed light on susceptibility genes and mechanisms for IS, we used the recently published GWAS data from the GIGASTROKE consortium. A TWAS analysis was also carried out by the GIGASTROKE group, which discovered 17 genes using prediction models based on GTEx v7 [16]. We found that 7/17 genes replicated with in our TWAS investigation in the brain, artery, and heart tissues with a strong correlation of TWAS effect-sizes (R = 0.73; see **Fig. S5**). Additionally, we identified 119 genes/proteins and demonstrated some strengths of our study. First, our reference panels for TWAS analysis were significantly larger than the GIGASTROKE study by leveraging GTEx v8 datasets [24], which allowed for more robust analysis and generalization of the findings. Second, our study used a diverse and extensive TWAS analysis, which included a multi-tissue mRNA/splicing/eRNA/protein transcriptome-wide association study (TWAS/spTWAS/eTWAS/PWAS). This diversity provides a broader range of biological mechanisms to understand IS etiology.

We note several limitations with our approach. First, our approach relies on using genetically predicted gene expression levels to identify genes whose expression associates with the genetic component of IS risk, which will require further functional studies for downstream validation. We note however, that our results were robust to choose TWAS approach (i.e., FUSION vs PrediXcan), statistical method (i.e., TWAS vs coloc), and eQTL reference panel (i.e., GTEx v7 vs GTEx v8). Second, we investigated the association between IS risk with diverse molecular contexts focusing on genetically predicted gene expression, protein abundance, splicing variation, and enhancer expression across multiple tissues when available. However, recent works have demonstrated improved association and colocalization power when investigating transcription factor binding, chromatin activity, and chromatin accessibility information [81,82], which may be potential mediators for IS risk, missed by our work. Despite these limitations, our results provide valuable insights into underlying molecular mechanisms and drug candidates for IS.

In conclusion, we highlight IS risk genes and proteins using the multi-tissue TWAS/spTWAS/eTWAS/PWAS approach. Moreover, we provided potential drug candidates for preventing IS. We believe that these findings could be used as valuable resources for understanding the underlying mechanisms and designing subsequent functional studies for IS treatment.

## Data Availability

All data produced in the present work are contained in the manuscript.

## Acknowledgements

This work was funded in part by the National Institutes of Health (NIH) under awards R01HG012133 and R01GM140287.

## URLs

The full results of TWAS, spTWAS, eTWAS, PWAS: https://github.com/mancusolab/stroke_twas

Iscemic stroke summary statistics (GIGASTROKE): https://www.ebi.ac.uk/gwas/studies/GCST90104540

GTEx v8 predictive models: http://gusevlab.org/projects/fusion/#gtex-v8-multi-tissue-expression

FUSION script: https://github.com/gusevlab/fusion_twas

INTERVAL predictive models: https://www.mancusolab.com/pwas

ARIC predictive models: http://nilanjanchatterjeelab.org/pwas/

LD blocks (GRCh19): https://bitbucket.org/nygcresearch/ldetect-data/src/master/EUR/

LD blocks (GRCh38): https://github.com/jmacdon/LDblocks_GRCh38

GWAS summary statistics (UK Biobank): http://www.nealelab.is/uk-biobank

