## Supplementary Figure for "Novel insight into the etiology of ischemic stroke gained by integrative transcriptome-wide association study"

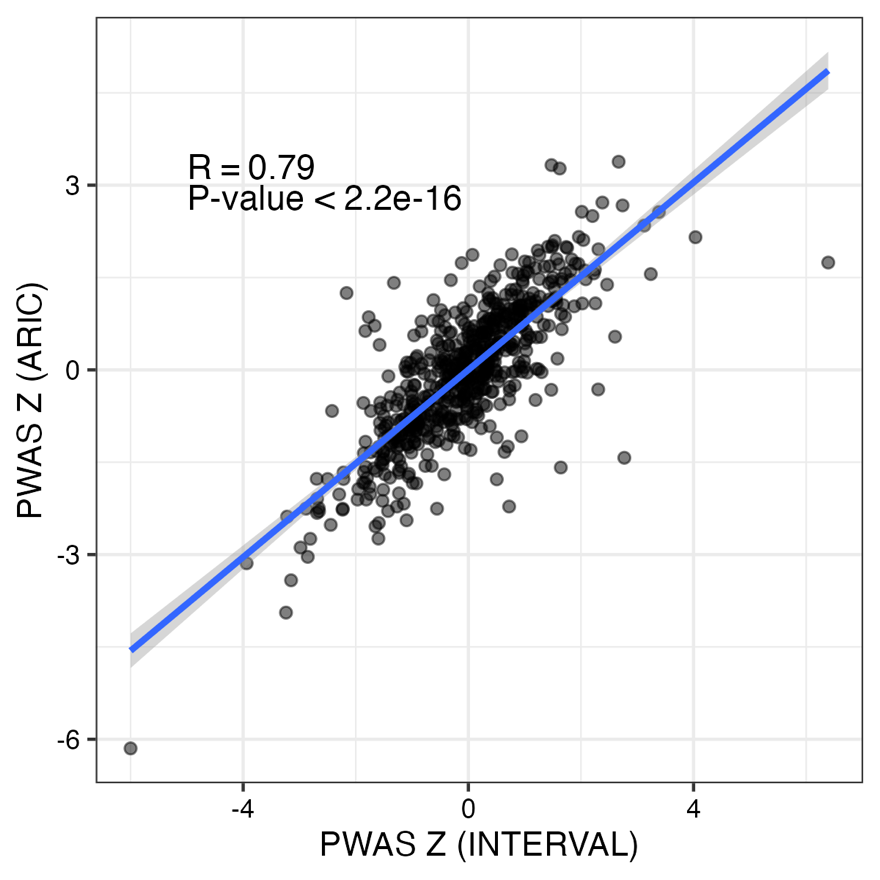

**Supplementary Figure 1. Comparison of PWAS Z scores from different reference panels.** Each point corresponds to a PWAS association from two different reference panels (e.g., INTERVAL and ARIC). Blue lines represent the best-fit regression line.

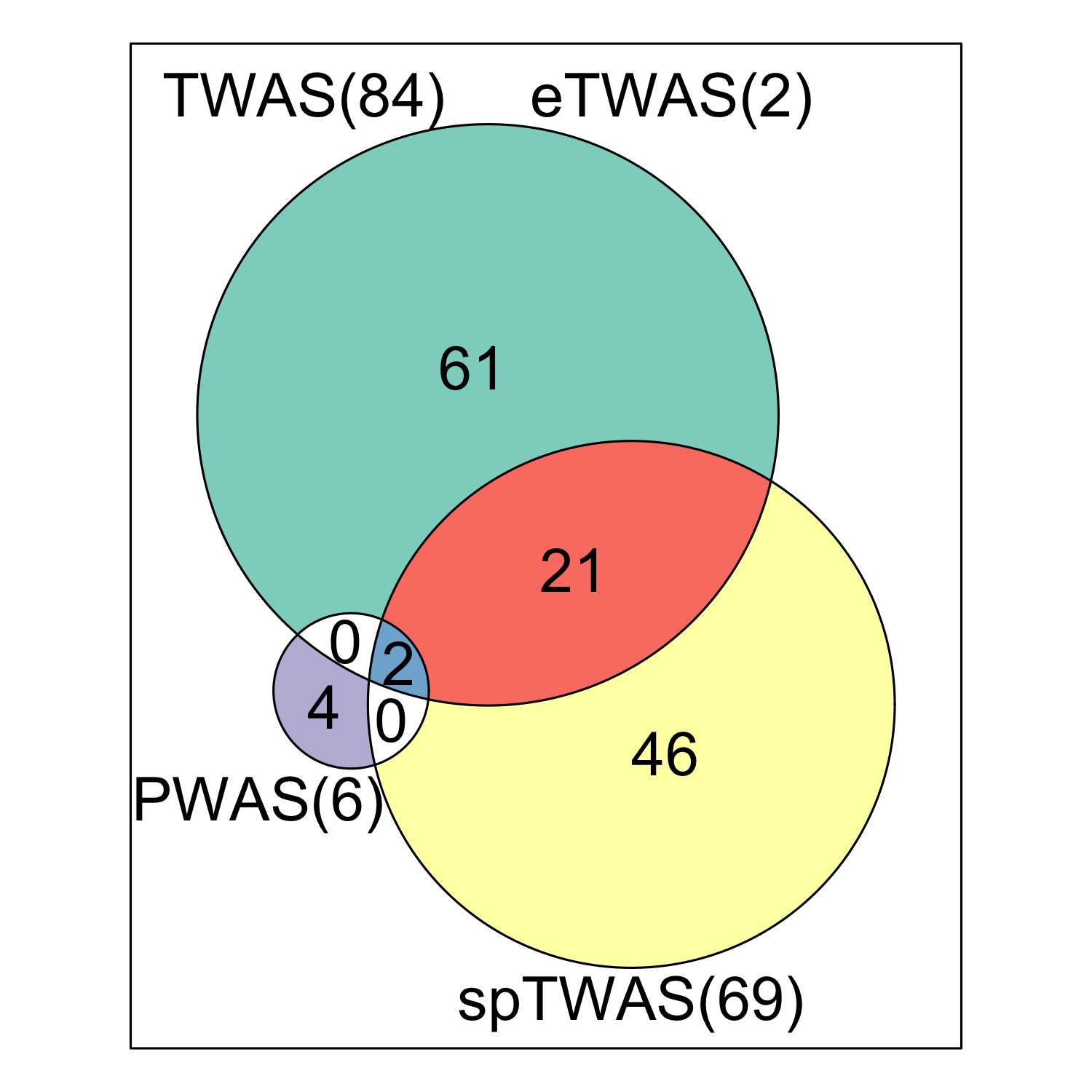

**Supplementary Figure 2. Results summary of TWAS/spTWAS/eTWAS/PWAS analysis.** A Venn diagram showing the summary of genes obtained from TWAS/spTWAS/eTWAS/PWAS.

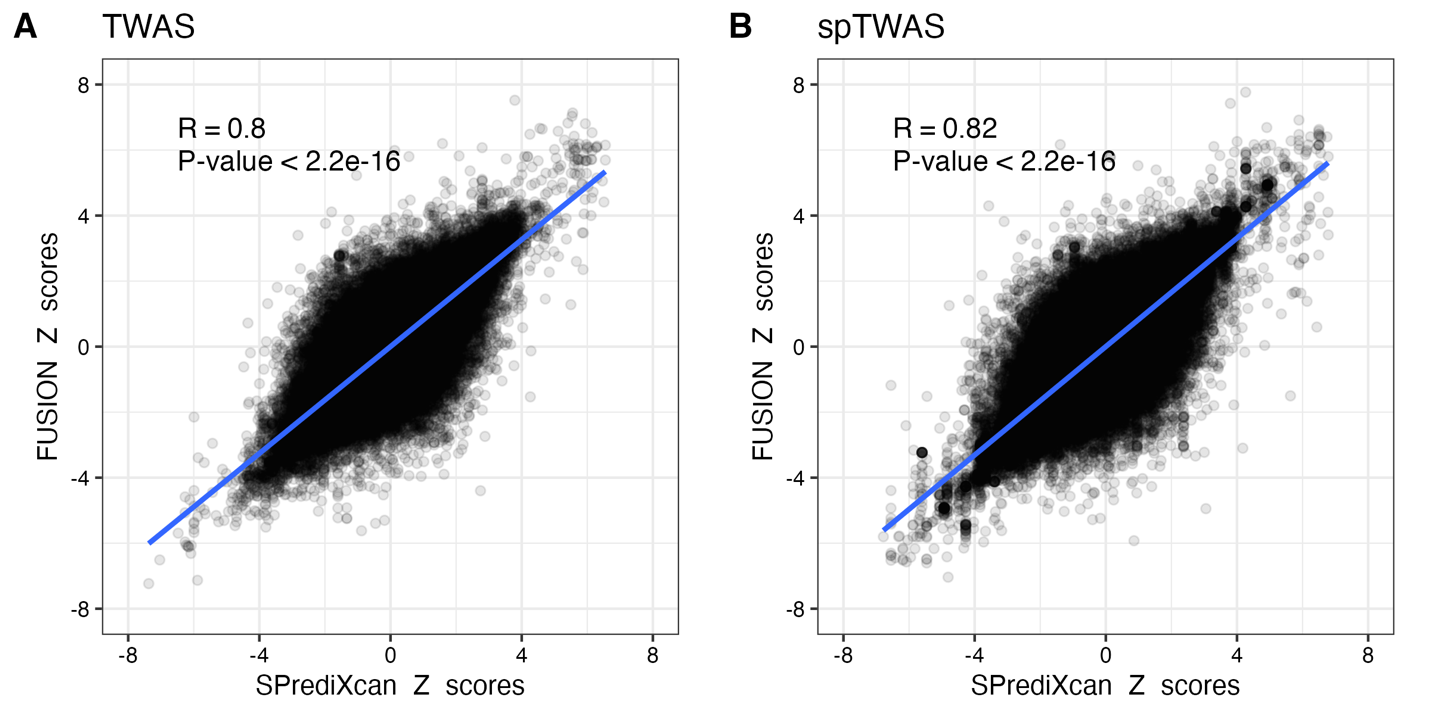

**Supplementary Figure 3. Comparison of Z scores from FUSION and S-PrediXcan.** Z scores from FUSION and S-PrediXcan using GTExv8 **(A)** eQTL data and **(B)** sQTL data. Each point corresponds to an association test between predicted gene expression or splicing with IS risk. Blue lines represent the best-fit regression line.

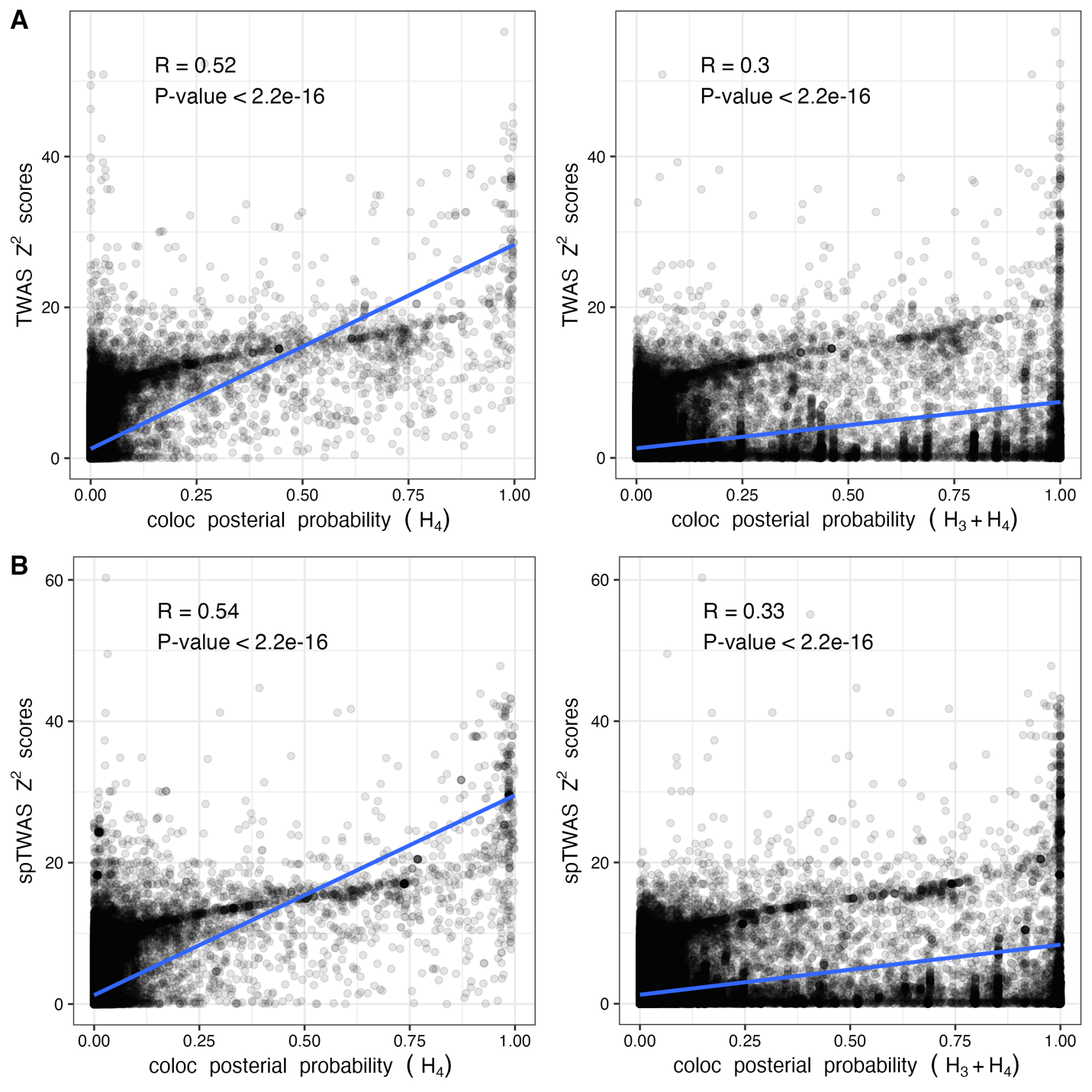

**Supplementary Figure 4. Relationship of TWAS and co-localization.** **(A)** The scatter plot shows the correlation between TWAS Z^2^ scores and co-localization posterior probability (PP) obtained from hypotheses *H_3_* and *H_4_*. **(B)** The scatter plot shows the correlation between spTWAS Z^2^ scores and co-localization posterior of *H_3_* and *H_4_*. *H_3_* is the PP of GWAS and eQTL signals are associated with different causal variants, and *H_4_* PP is the PP of GWAS and eQTL signals are associated and share a single causal variant. Blue lines represent the best-fit regression line.

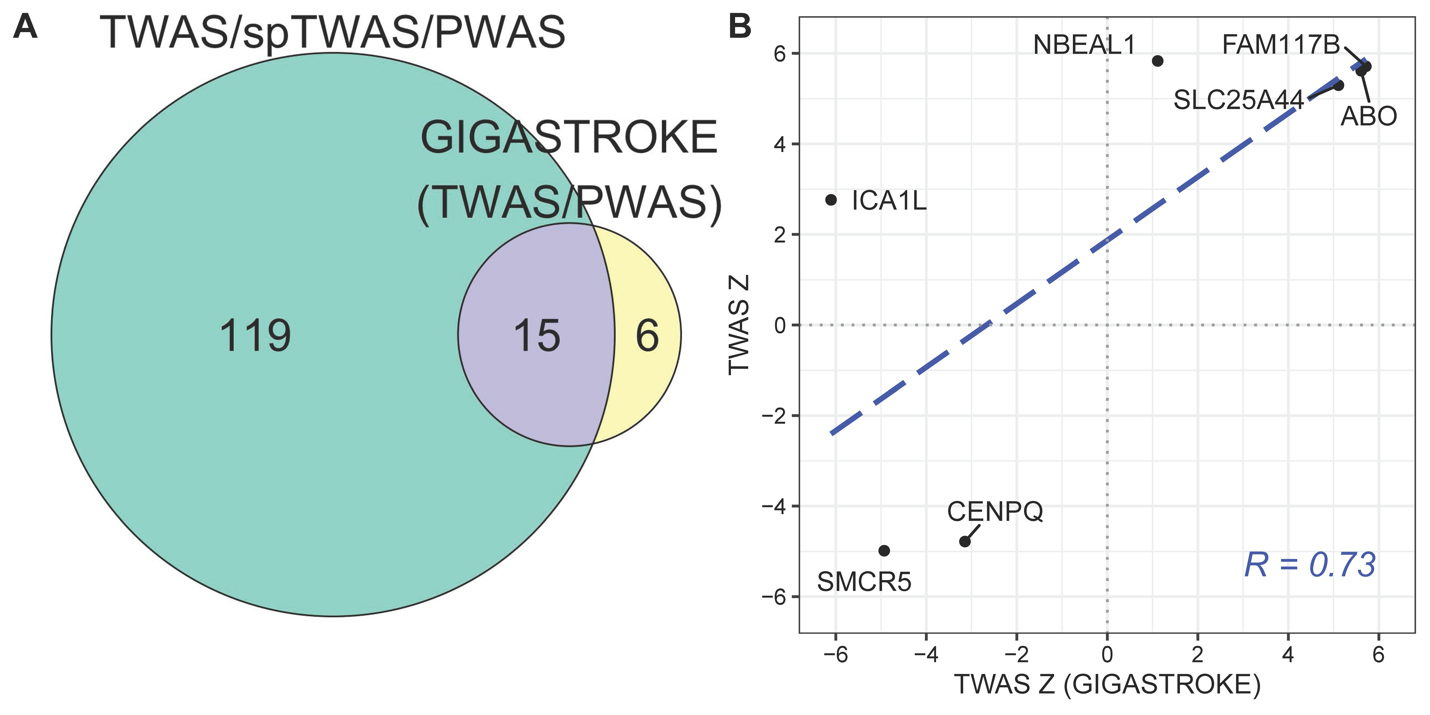

**Supplementary Figure 5. Comparison of previous findings. (A)** A Venn diagram showing the overlap genes/proteins between TWAS/PWAS from the GIGASTROKE study and TWAS/spTWAS/PWAS analysis in this study. **(B)** Comparison of TWAS Z scores from GIGASTROKE study. Each point corresponds to a TWAS association between TWAS from the GIGASTROKE study and TWAS analysis in this study. Blue lines represent the best-fit regression line.

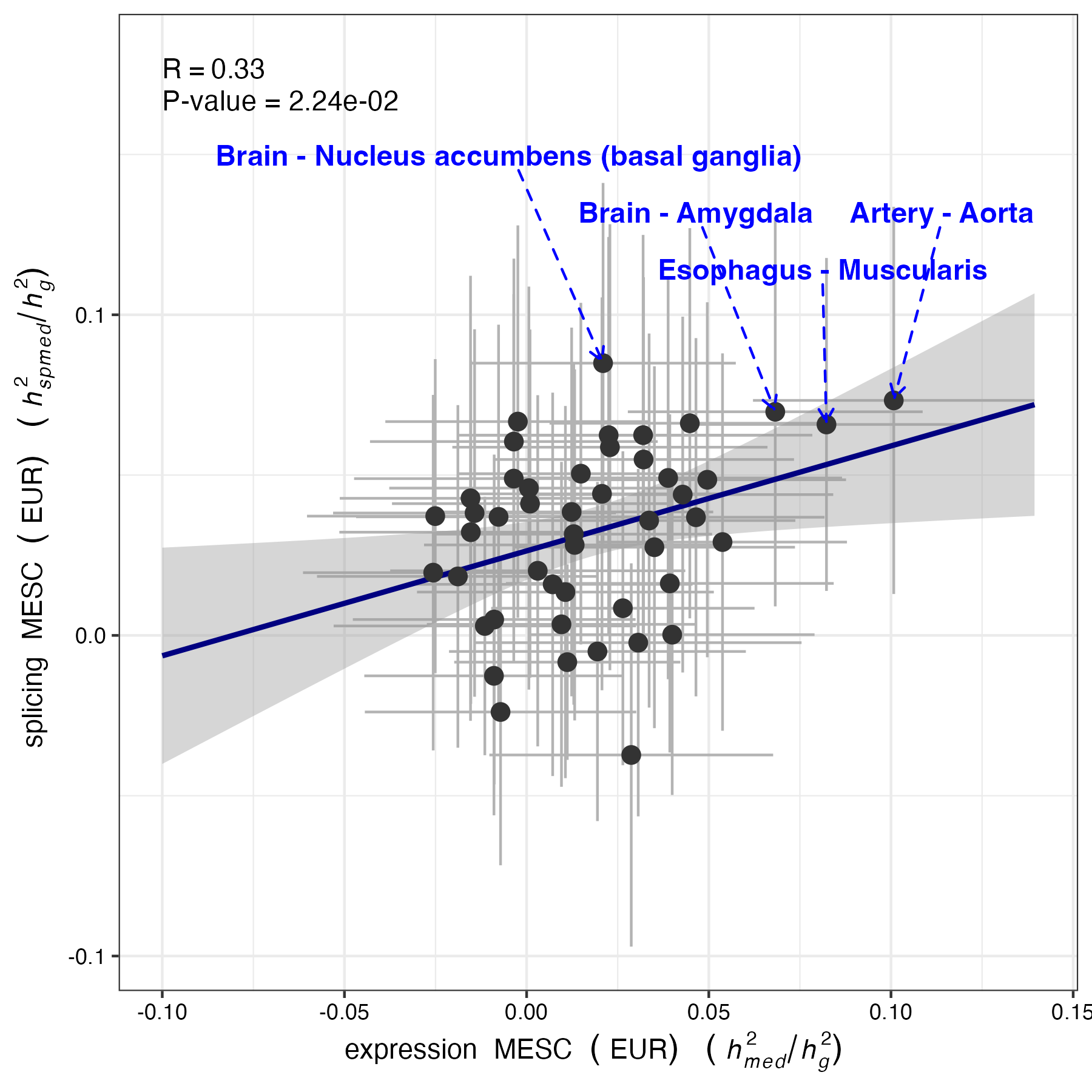

**Supplementary Figure 6. Relationship between expression MESC and splicing MESC**. Expression MESC $(h_{med}^{2}/h_{g}^{2})$ and splicing MESC $(h_{spmed}^{2}/h_{g}^{2})$ estimates from expression and alternative splicing scores estimated in each of 48 individual GTEx tissues, respectively. Each error bar indicates jackknife standard errors. Blue lines represent the best-fit regression line.

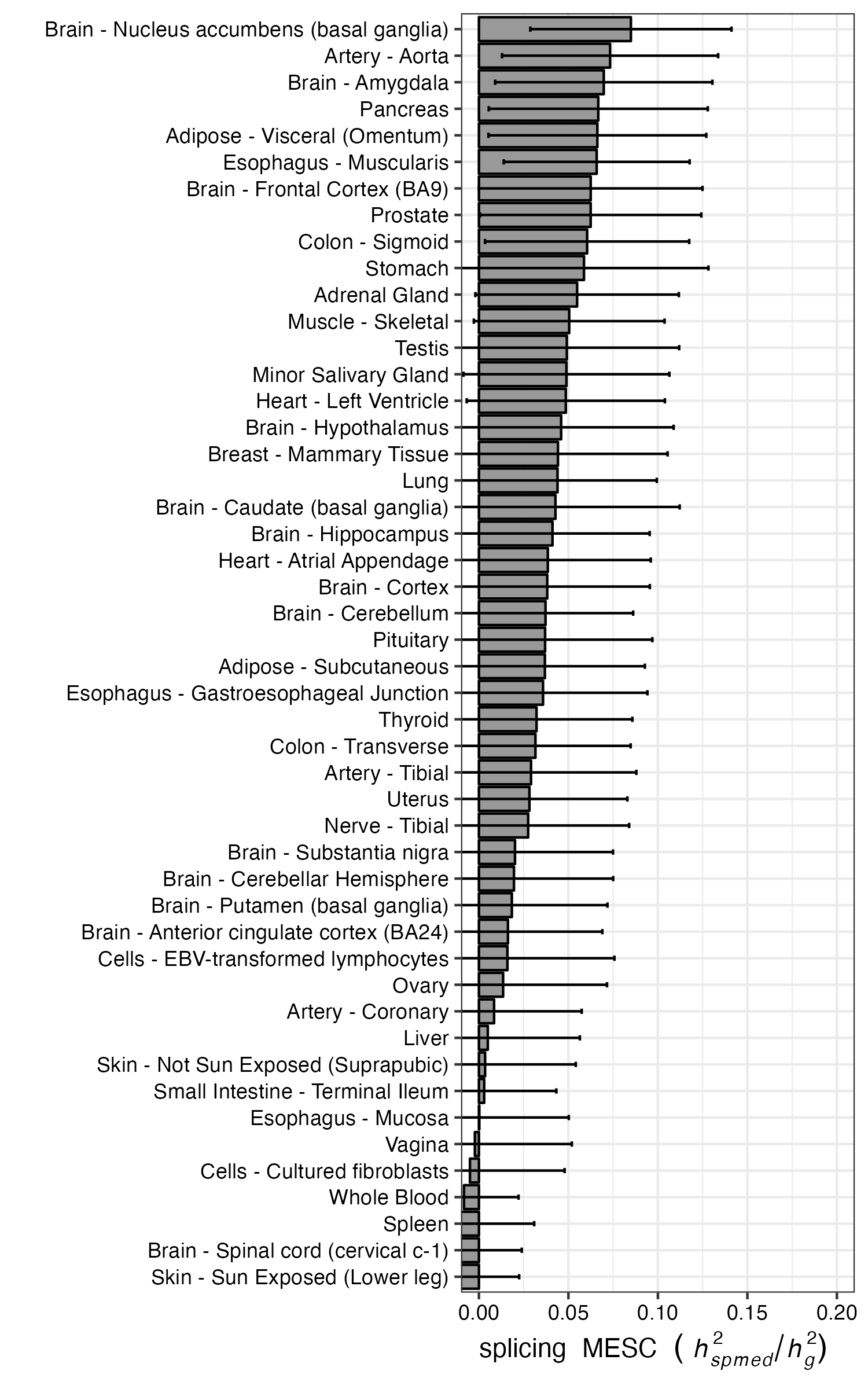

**Supplementary Figure 7. The proportion of heritability mediated by alternative splicing.** The bar plots correspond to the estimated splicing MESC ($h_{spmed}^{2}/h_{g}^{2}$) in each tissue panel in GTEx v8. Each error bar indicates jackknife standard errors.

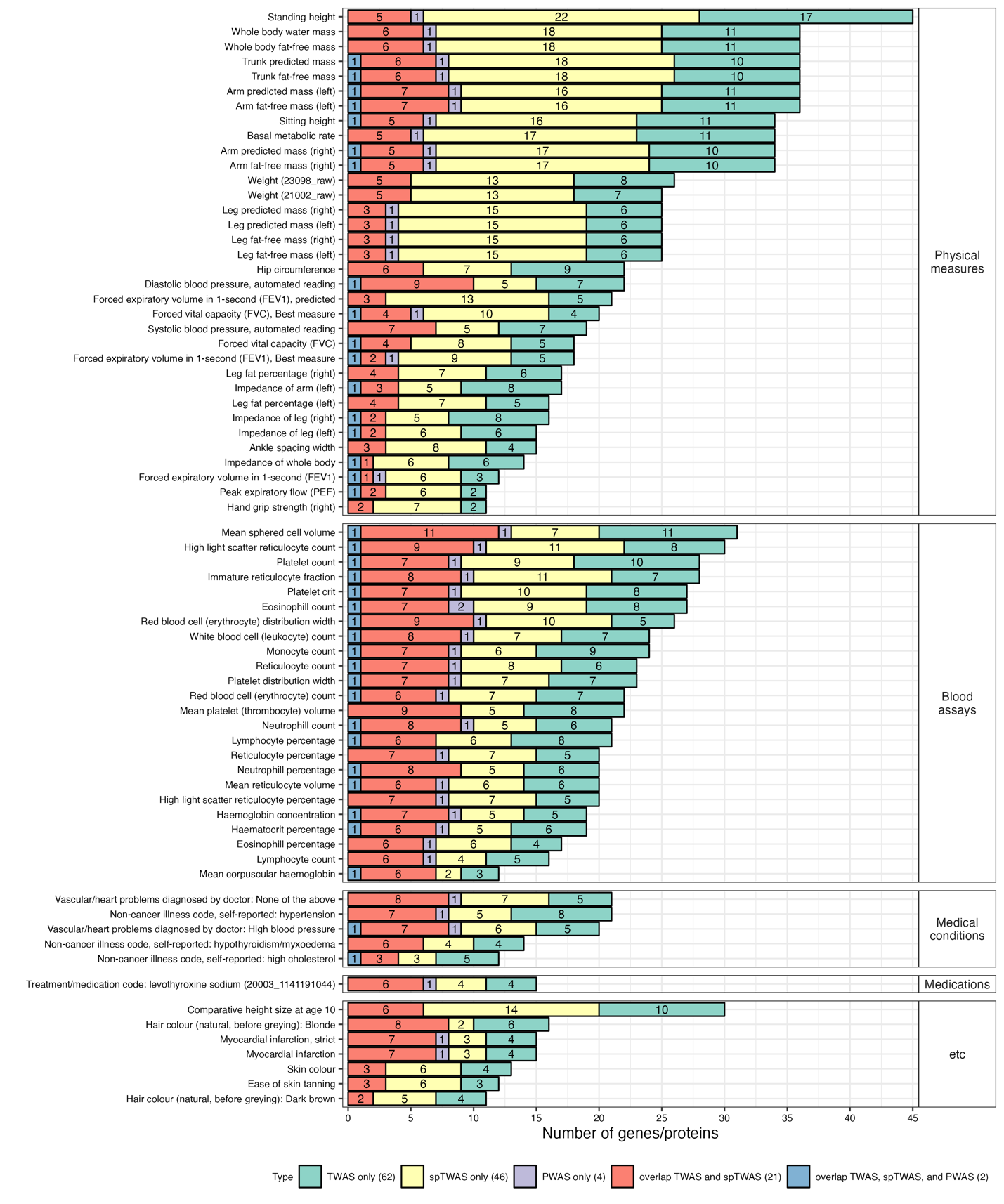

**Supplementary Figure 8. Phenome-wide association study.** The Bar plot represents a number of overlap genes between IS TWAS/spTWAS/PWAS and different phenotypes retrieved from PhenomeXcan.

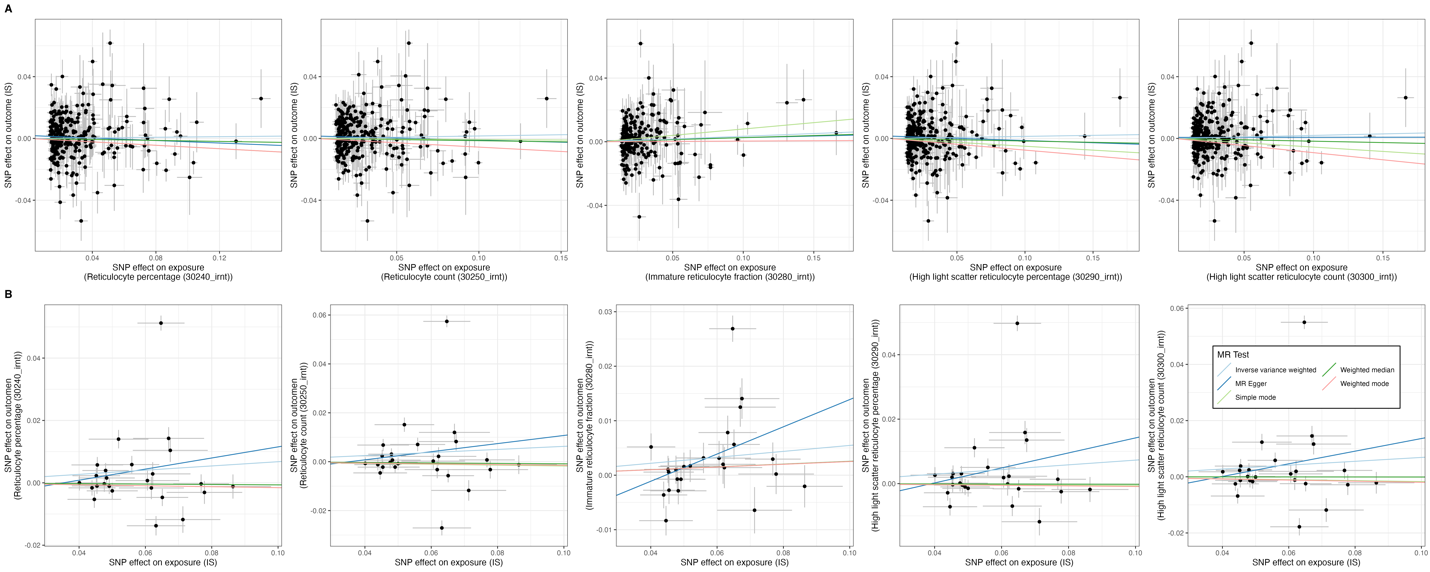

**Supplementary Figure 9. Mendelian randomization (MR) between IS and reticulocyte traits.** (A) MR was carried out using independent genetic instruments associated with five reticulocyte traits. (B) MR was carried out using independent genetic instruments associated with IS. Each point represents the SNP effects on IS and reticulocyte traits.

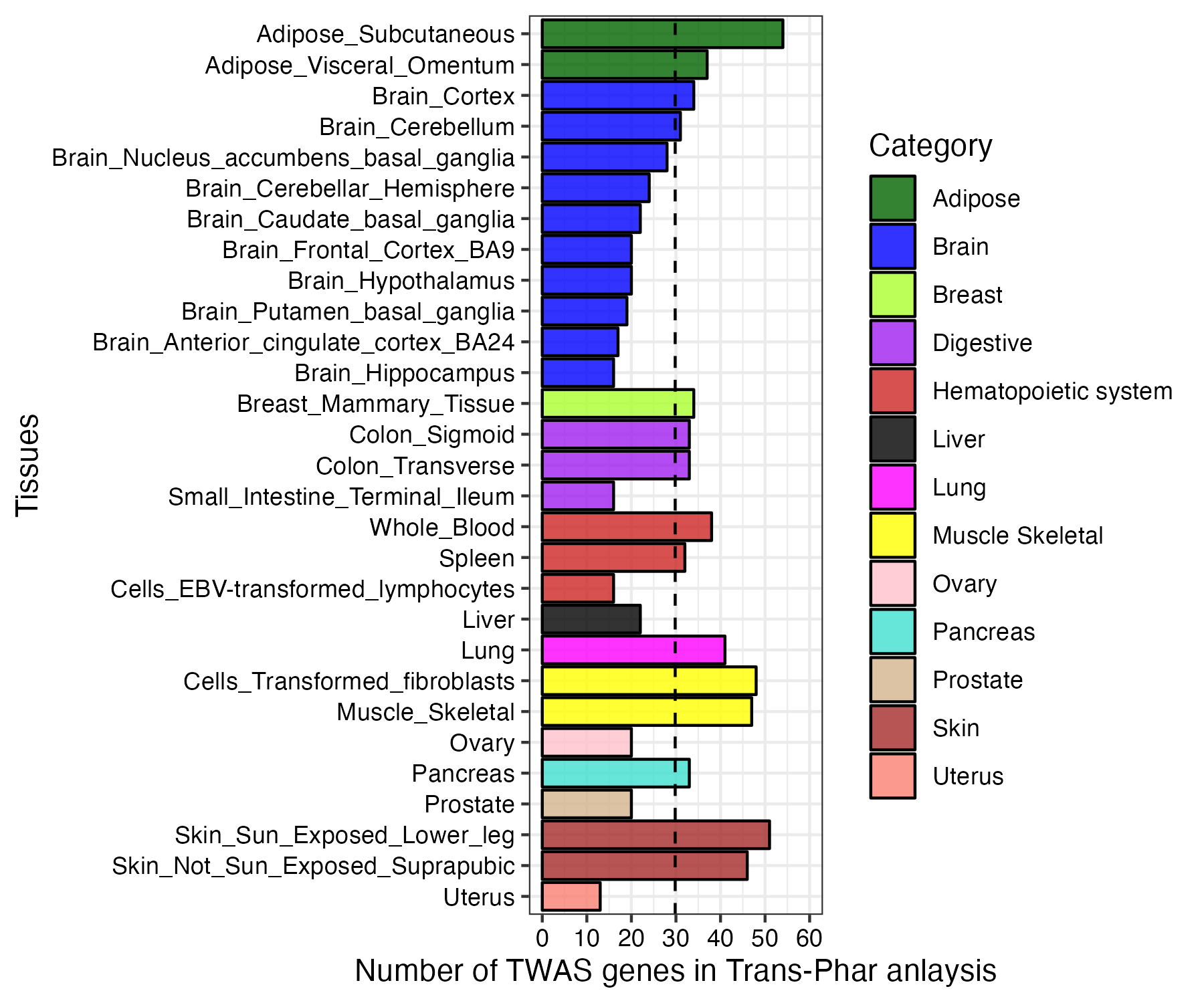

**Supplementary Figure 10. The Number of TWAS/spTWAS/PWAS genes in Trans-Phar analysis in each tissue.** The color bars indicate the tissue category of the GTEx tissue. The dotted line represents an average value of the number of TWAS/spTWAS/PWAS genes in Trans-Phar analysis.

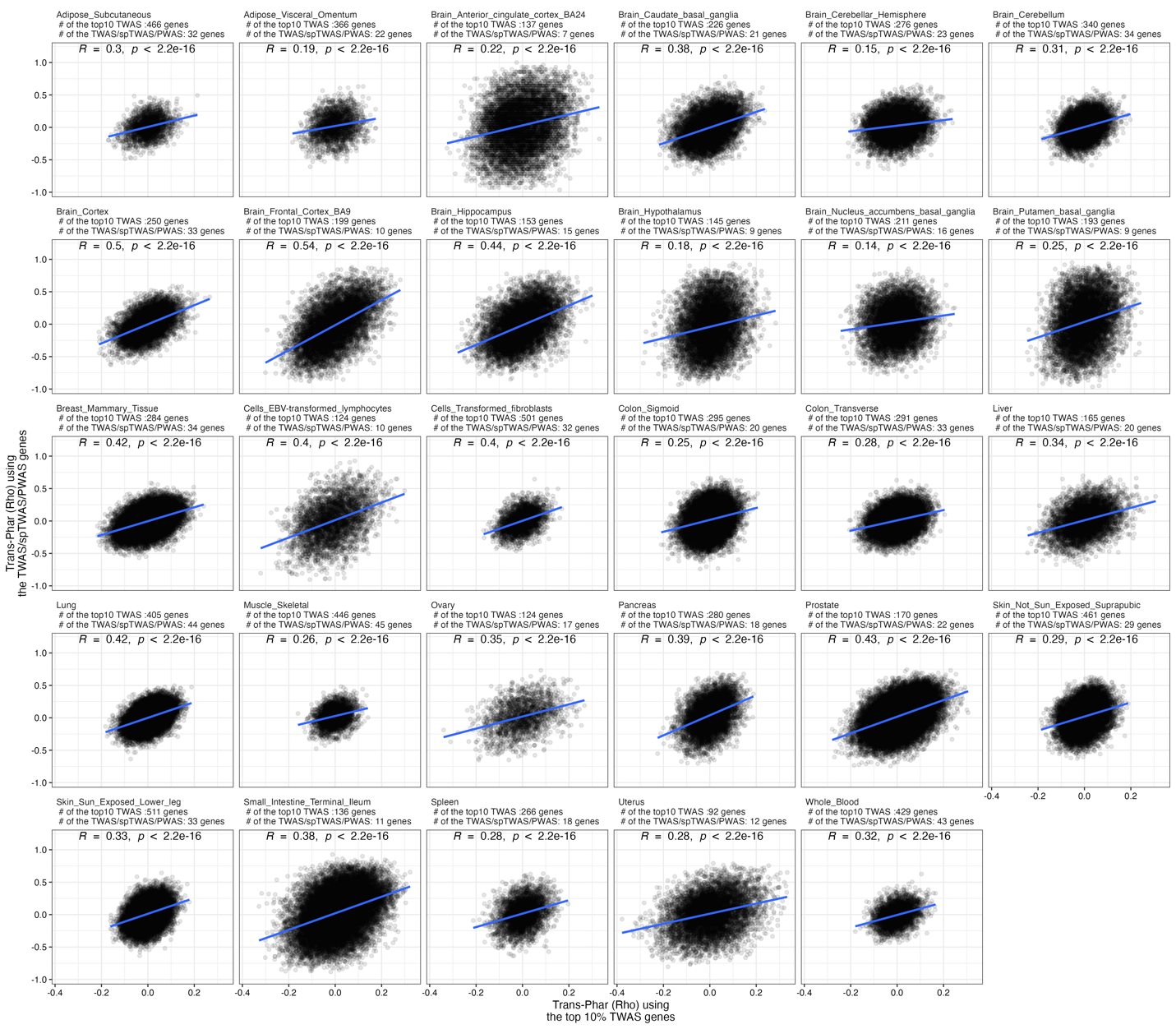

**Supplementary Figure 11. Relationship between Trans-Phar results from the top 10% TWAS genes and the 134 TWAS/spTWAS/PWAS genes.** The number of genes used in the analysis is indicated at the top of each tissue result. Spearman's rank correlation coefficient and P-value are indicated in each tissue. Blue lines represent the best-fit regression line.

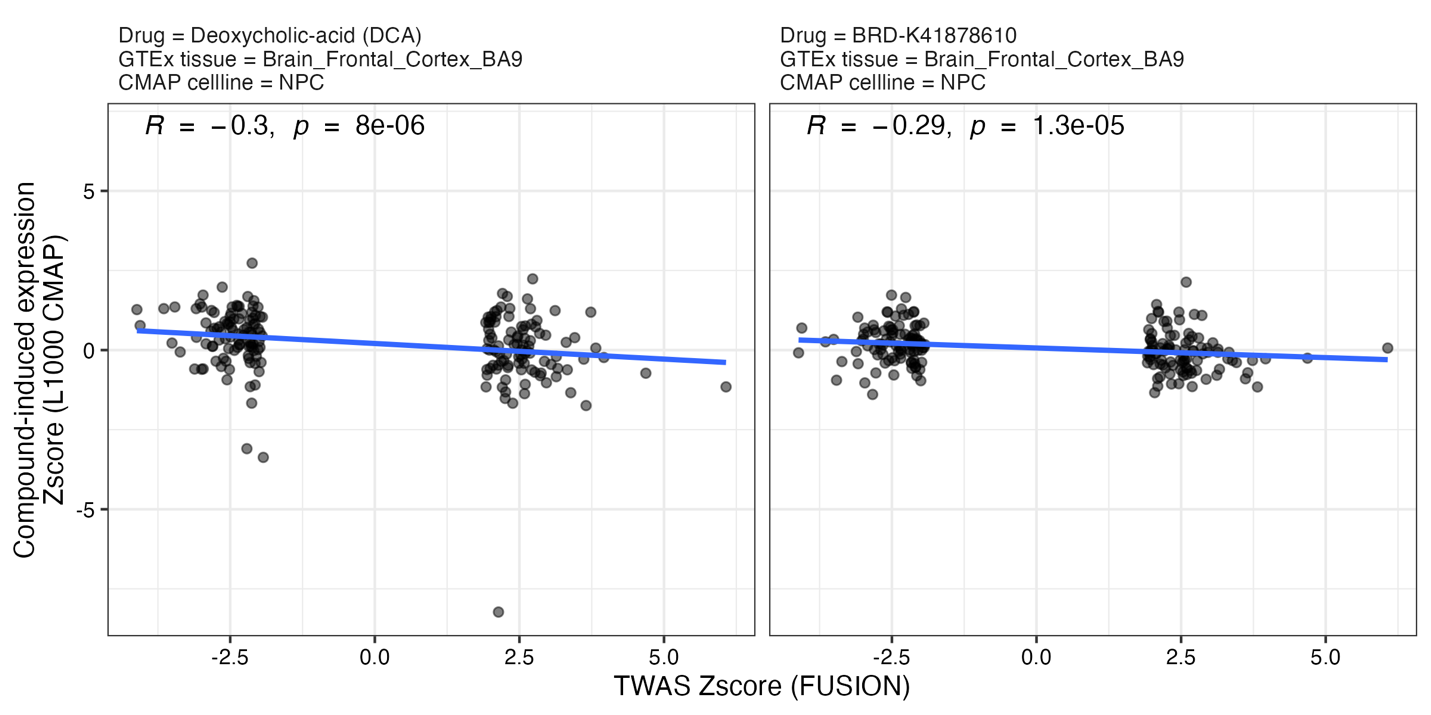

**Supplementary Figure 12. The significant TWAS-compound linkages from Trans-Phar.** The drug name, GTEx tissue, and CMAP cell line are indicated at the top of each drug result. Spearman's rank correlation coefficient and P-value are indicated in each tissue. Blue lines represent the best fit regression line.

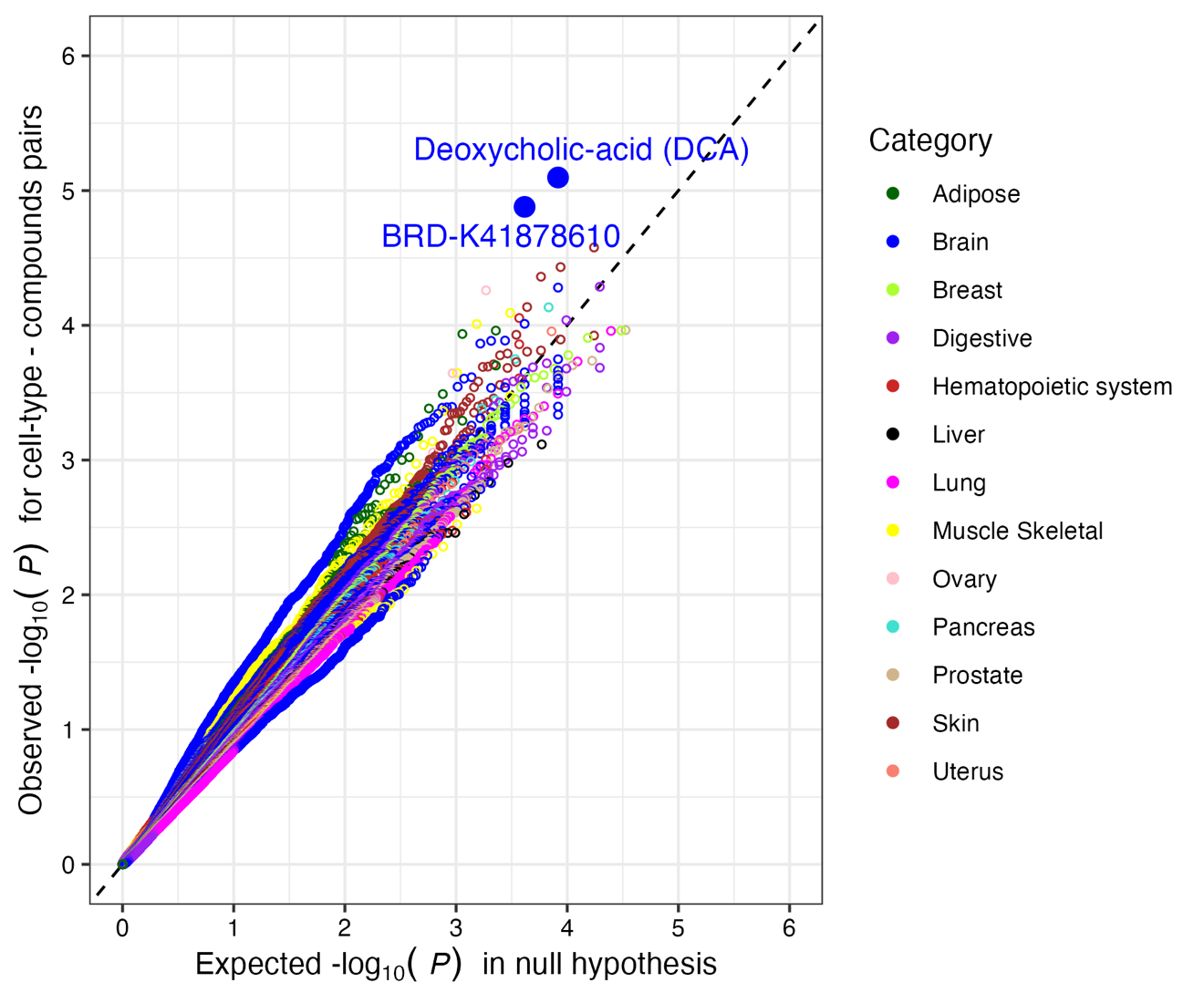

**Supplementary Figure 13. Q–Q plots for TWAS-compound linkages.** The x-axis corresponds to an expected distribution of P-values under the null hypothesis. The y-axis corresponds to an observed distribution of P-values per tissue or cell-type.
